# Long term association between frailty and survival in patients with pleural disease: a cohort study protocol

**DOI:** 10.1101/2023.02.06.23285492

**Authors:** E Barton, B Carter, A Verduri, J Hughes, J Hewitt, NA Maskell

**Affiliations:** Academic Respiratory Unit, School of Clinical Sciences, University of Bristol, UK; Department of Biostatistics and Health Informatics, Institute of Psychiatry, Psychology and Neuroscience, Kings College London, UK; Respiratory Unit, Department of Surgical and Medical Sciences, University of Modena and Reggio Emilia, Policlinico Modena, Italy; Department of Population Medicine, Cardiff University Cardiff, UK

**Keywords:** Pleural disease, frailty, survival

## Abstract

**Introduction:** As the population ages, frailty is becoming more common and understanding how frailty impacts on patient outcomes has become an integral part of clinical care. To date, there is no evidence available on the relationship between frailty and patient outcomes in pleural disease. In this study we explore the relationship between frailty and both malignant and non-malignant pleural disease using the modified frailty index (mFI).

**Methods and Analysis:** Outpatients with pleural disease will be identified from a prospectively collected single-centre UK database and their data and notes retrospectively analysed. An mFI will be calculated for each patient and the correlation between their frailty index, final diagnosis and mortality analysed.

**Dissemination:** Study findings will be disseminated by publication in an appropriate journal and presentations at Respiratory and/or Geriatric medicine meetings.

## Introduction

Frailty is a clinical state of increased vulnerability to external stressors such as illness, in which multiple body systems gradually lose their reserves.^1^ Frailty assessment and intervention represents a huge challenge to the healthcare system worldwide. Frailty can impact on every aspect of patient care and its assessment may improve patient management and clinical outcomes.

Th UK’s population is aging; in 2016, 18% of the UK population were aged >65, an increase of 2.2% over the preceding 25 years. This is projected to increase to 26% in the next 25 years.^2^ That there is an association between age and frailty is not in dispute, but age alone is at best a surrogate marker for the poor physiological reserve seen in frail patients. Frailty is known to be associated with adverse health outcomes including increased mortality risk^3^, but defining and assessing frailty can be challenging.^4^

The most commonly described approaches to identify frailty are the frailty index (FI), which is a quantification of the cumulative burden of health deficits^5^, or the phenotype model based on domains such as weakness, slowness and low physical activity.^6^ The FI is historically based on a cumulative burden of 70 individual deficits. Therefore shorter versions, with more clinical utility, have been developed, such as the modified Frailty Index (mFI).^7^ Studies have shown that patients with higher frailty scores have higher mortality and a higher risk of post-operative complications, delirium and institutionalisation.^8–11^

Pleural disease is common; an estimated 360/100,000 people are diagnosed with pleural disease each year.^12^ Approximately a quarter of pleural effusions are attributed to malignancy and a quarter to cardiac failure, both common conditions in the aged.^13^ Most data on frailty in pulmonary disease are limited to those with Chronic Obstructive Pulmonary Disease (COPD) and interstitial lung disease (ILD) and there is currently no data available on the relationship between frailty and patient outcomes in pleural disease. In a cohort of patients with such a high incidence of malignancy, in whom life expectancy is likely limited and quality of life should be prioritised, understanding the relationship between frailty and outcomes, including mortality, could be highly beneficial to assess who may be a suitable candidate for aggressive investigation and management.

In this study, we explore the relationship between frailty and both malignant and non-malignant pleural disease and evaluate the utility of the mFI in assessing this cohort of patients.

### Study aims

Our main aim is to assess the prevalence of frailty in patients with both malignant and non-malignant pleural disease. We hypothesise that the presence of frailty is associated with increased mortality in patients with pleural disease.

## Methods

### Study design

Since 2008, North Bristol NHS Trust has collected one of the largest repositories of pleural fluid samples and data in the world, including over 1500 patients with pleural disease. Consecutive patients aged 18 years or older presenting to North Bristol NHS Trust’s tertiary pleural service with radiographically confirmed undiagnosed pleural effusions were recruited to this prospective observational study and underwent a comprehensive diagnostic evaluation to identify the underlying aetiology of their pleural effusion. Data regarding demographics and comorbidities were systematically collected at recruitment. The diagnosis of a comorbidity was confirmed by patient’s medication list and medical record. 12 months following presentation, a final diagnosis was recorded independently by two respiratory consultants. Samples and data from this cohort of patients have been utilised in multiple studies previously, to improve the diagnostic and management pathway of patients with pleural disease.^14–17^

This is a retrospective analysis of data from outpatients recruited to this single-centre database between March 2008 and December 2020.

### Procedures for pleural fluid analysis

Pleural fluid samples collected at presentation and enrolment in the study underwent biochemical and cytological analysis. If required, histological samples were also collected either by radiologically guided percutaneous pleural biopsy, or under direct vision at medical thoracoscopy or video-assisted thoracoscopic surgery (VATS) in cases of suspected malignant pleural effusion.

### Data extraction

Patients presenting as outpatients to the pleural service between 2008 and 2020 will be identified from the study database and their records retrospectively analysed and cross-referenced with their available electronic patient record. Patients on whom no mortality or comorbidity data is available will be excluded. A modified frailty index (mFI) for each patient will be calculated based on these data.

### Study measurements

The following data will be extracted from patients’ electronic patient record:

❖Age

❖Sex

❖Date of presentation

❖Asbestos exposure

❖Performance status (WHO performance status or Karnofsky performance status)

❖Functional dependence

❖Co-morbidities – specifically diagnoses of Chronic Obstructive Pulmonary Disease (COPD), treated hypertension, diabetes and chronic heart failure at presentation

❖Date of death (if applicable)

❖Survival at 12 months from presentation

❖Final diagnosis at 12 months

### Frailty assessment

For this study, frailty was assessed using the modified FI (mFI), a validated score that has been specifically designed for assessment using clinical datasets and electronic health records. The modified frailty index (mFI) was developed by the American College of Surgeons National Surgical Quality Improvement Program (ACS NSQIP); it produces a score based on 5 factors, which has been demonstrated to strongly predict mortality and post-operative complications.^18^ The mFI includes five items: chronic heart failure (CHF); Chronic Obstructive Pulmonary Disease (COPD); diabetes mellitus (DM); being on treatment for hypertension (HPT) and functional dependence as the component deficits. The mFI is based on the cumulative deficit model where the more comorbidities present, the higher the frailty index (FI), derived as the number of deficits divided by the number of deficits assessed. This gives a mFI range of 0-1, with each contributing domain assigned a score of 0.2. For analyses, mFI was categorised as not frail (mFI <0.4) and frail (mFI ≥0.4). CHF, COPD, DM, and treated HPT were identified using electronic clinical records; functional dependence was assessed using the Karnofsky Performance Status score (cut off score for functional dependence = 60), the WHO Performance Status score (cut off score for functional dependence = 3), and electronic clinical records.

## Outcomes

### Primary outcome

❖Time to all-cause mortality from date of presentation.

## Statistical analysis

This protocol and analysis have been drafted by a statistician fully blinded to the outcome data (BC) following King’s College London Clinical Trials Unit (KCTU) Standard operating procedures on drafting a Statistical Analysis Plan (SAP).

### Primary analysis

The time to all-cause mortality will be fitted to compare those classified as frail versus not frail visually using a Kaplan Meier plot with associated at risk table and log-rank test. Data will be analysed using a multivariable Cox baseline proportional hazards regression to all-cause mortality, adjusting for: age at diagnosis (Under 64, 65 to 79, 80 or older); underlying aetiology; suspected asbestos exposure status, performance status, malignancy status and frailty (using the mFI). We will present the crude hazard ratio (HR) and adjusted HR (aHR) with associated 95% confidence intervals and p-values. The baseline proportional hazard assumption will be assessed visually using log-log plots.

Missing data will be explored for pattern missingness. Variables with greater than 5% of missingness may be imputed following Jacobson et al 2017.^19^

### Subgroup analyses

We will carry out a subgroup analyses for age groups and final diagnosis and will present the effect of being frail compared to not being frail for each subgroup.

## Discussion

This study will provide the first observational data on the association between frailty and a range of standard clinical outcomes in people living with frailty. These results are expected to be presented at relevant medical conferences published in a peer reviewed journal in the second half of 2023.

## Signatures

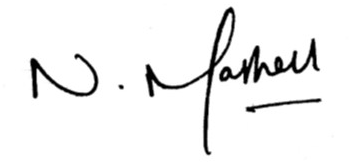

Professor Nick Maskell

Professor of Respiratory Medicine

Academic Respiratory Unit, Bristol

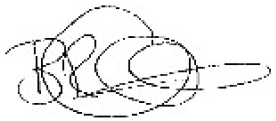

Dr Ben Carter

Reader in Medical Statistics

(Study Statistician)

King’s College London

## Data Availability

All data produced in the present study will be contained in the manuscript.

## References

1. Clegg A, Young J, Iliffe S, Rikkert MO, Rockwood K. Frailty in elderly people. The Lancet 2013;381(9868):752–62.

2. Storey A. Living longer: how our population is changing and why it matters [Internet]. 2018 [cited 2023 Jan 10];Available from: https://www.ons.gov.uk/peoplepopulationandcommunity/birthsdeathsandmarriages/ageing/articles/livinglongerhowourpopulationischangingandwhyitmatters/2018-08-13

3. García-González JJ, García-Peña C, Franco-Marina F, Gutiérrez-Robledo LM. A frailty index to predict the mortality risk in a population of senior mexican adults. BMC Geriatr 2009;9(1):47.

4. Bergman H, Ferrucci L, Guralnik J, et al. Frailty: An Emerging Research and Clinical Paradigm--Issues and Controversies. J Gerontol A Biol Sci Med Sci 2007;62(7):731–7.

5. Song X, Mitnitski A, Rockwood K. Prevalence and 10-Year Outcomes of Frailty in Older Adults in Relation to Deficit Accumulation. J Am Geriatr Soc 2010;58(4):681–7.

6. Fried LP, Tangen CM, Walston J, et al. Frailty in Older Adults: Evidence for a Phenotype. J Gerontol A Biol Sci Med Sci 2001;56(3):M146–57.

7. Carter B, Keevil VL, Anand A, et al. The Prognostic and Discriminatory Utility of the Clinical Frailty Scale and Modified Frailty Index Compared to Age. Geriatrics 2022;7(5):87.

8. Jones DM, Song X, Rockwood K. Operationalizing a Frailty Index from a Standardized Comprehensive Geriatric Assessment. J Am Geriatr Soc 2004;52(11):1929–33.

9. Makary MA, Segev DL, Pronovost PJ, et al. Frailty as a Predictor of Surgical Outcomes in Older Patients. J Am Coll Surg 2010;210(6):901–8.

10. Lee DH, Buth KJ, Martin B-J, Yip AM, Hirsch GM. Frail Patients Are at Increased Risk for Mortality and Prolonged Institutional Care After Cardiac Surgery. Circulation 2010;121(8):973–8.

11. Farhat JS, Velanovich V, Falvo AJ, et al. Are the frail destined to fail? Frailty index as predictor of surgical morbidity and mortality in the elderly. Journal of Trauma and Acute Care Surgery 2012;72(6):1526–31.

12. Bodtger U, Halifax R J. Epidemiology: why is pleural disease becoming more common? In: Maskell NA, Laursen CB, Lee YCG, Rahman NM, editors. Pleural Disease. European Respiratory Society; 2020. p. 1–12.

13. Porcel JM, Esquerda A, Vives M, Bielsa S. Etiology of Pleural Effusions: Analysis of More Than 3,000 Consecutive Thoracenteses. Archivos de Bronconeumología (English Edition) 2014;50(5):161–5.

14. Dixon G, Bhatnagar R, Zahan-Evans N, et al. A Prospective Study to Evaluate a Diagnostic Algorithm for the Use of Fluid Lymphocyte Subset Analysis in Undiagnosed Unilateral Pleural Effusions. Respiration 2018;95(2):98–105.

15. Bibby AC, Halford P, de Fonseka D, Morley AJ, Smith S, Maskell NA. The Prevalence and Clinical Relevance of Nonexpandable Lung in Malignant Pleural Mesothelioma. A Prospective, Single-Center Cohort Study of 229 Patients. Ann Am Thorac Soc 2019;16(10):1273–9.

16. Arnold DT, Hamilton FW, Elvers KT, et al. Pleural Fluid suPAR Levels Predict the Need for Invasive Management in Parapneumonic Effusions. Am J Respir Crit Care Med 2020;201(12):1545–53.

17. de Fonseka D, Arnold DT, Morley AJ, et al. Lymphocyte predominance in blood, pleural fluid, and tumour stroma; a prognostic marker in pleural mesothelioma. BMC Pulm Med 2022;22(1):173.

18. Subramaniam S, Aalberg JJ, Soriano RP, Divino CM. New 5-Factor Modified Frailty Index Using American College of Surgeons NSQIP Data. J Am Coll Surg 2018;226(2):173–181e8.

19. Jakobsen JC, Gluud C, Wetterslev J, Winkel P. When and how should multiple imputation be used for handling missing data in randomised clinical trials – a practical guide with flowcharts. BMC Med Res Methodol 2017;17(1):162.

